# Adiponectin/leptin ratio - a marker of insulin sensitivity in pre-eclampsia and fetal growth

**DOI:** 10.1101/2022.10.13.22281056

**Authors:** Victoria Elizabeth de Knegt, Paula L. Hedley, Anna K. Eltvedt, Sophie Placing, Karen Wøjdemann, Anne-Cathrine Shalmi, Line Rode, Jørgen Kanters, Karin Sundberg, Ann Tabor, Ulrik Lausten-Thomsen, Michael Christiansen

**Affiliations:** Department for Congenital Disorders, Statens Serum Institut, Copenhagen, Denmark; Department of Paediatrics, University Hospital Slagelse, Slagelse, Denmark; Brazen Bio, Los Angeles, California, USA; Department of Gynecology and Obstetrics, Bornholm Hospital, Rønne, Denmark; Department of Obstetrics, Hillerød Hospital, Hillerød, Denmark; Department of Clinical Biochemistry, Copenhagen University Hospital Rigshospitalet, Glostrup, Denmark; Department of Biomedical Sciences, University of Copenhagen, Copenhagen, Denmark; Center of Fetal Medicine, Department of Obstetrics, Copenhagen University Hospital Rigshospitalet, Copenhagen, Denmark; Department of Clinical Medicine, Faculty of Medical and Health Sciences, University of Copenhagen, Copenhagen, Denmark; Department of Neonatology, University Hospital Rigshospitalet, Copenhagen, Denmark

**Keywords:** adipocytokine, adiponectin, birth weight, fetal growth restriction, leptin, metabolic syndrome, placenta

## Abstract

The serum adiponectin-leptin ratio (A/L ratio) is a surrogate marker of insulin sensitivity. Pre-eclampsia (PE) is associated with maternal metabolic syndrome and occasionally impaired fetal growth. We assessed whether the A/L ratio in first-trimester maternal serum was associated with PE and/or birth weight. Adiponectin and leptin were quantitated in first-trimester blood samples (gestational week 10^+3^-13^+6^) from 126 women who later developed PE with proteinuria, (98 mild PE; 21 severe PE; 7 HELLP syndrome), and 297 controls, recruited from the Copenhagen First-Trimester Screening Study. The A/L ratio was reduced in PE pregnancies, median 0.17 (IQR: 0.12-0.27) compared to controls, median 0.32 (IQR: 0.19-0.62), (p<0.001). A multiple logistic regression showed that PE was negatively associated with A/L ratio independent of maternal BMI (odds ratio = 0.08, 95% CI = 0.0322 to 0.214). Adiponectin (AUC = 0.632) and PAPP-A (AUC = 0.605) were negatively, and leptin (AUC = 0.712) was positively associated with PE. However, the A/L ratio was a better predictor of PE (AUC = 0.737). No significant association was found between A/L ratio and clinical severity of pre-eclampsia or preterm birth. PE was associated with significantly lower relative birth weight, (p<0.001). A significant negative correlation was found between relative birth weight and A/L ratio in controls but not in PE pregnancies, (*β* = −0.144, 95% CI = −9.944 to −0.093), independent of maternal BMI. After correction for maternal BMI, leptin was significantly associated with relative birth weight, (*β* = 0.197, 95 % CI = 2.361 to 14.353), while adiponectin was not significantly associated. Our findings suggest that an impairment of the A/L ratio (as seen in metabolic syndrome) in first-trimester is characteristic of PE, while aberrant fetal growth in PE is not dependent on insulin sensitivity but rather on leptin associated pathways.

## Background

Pre-eclampsia (PE) is a serious pregnancy complication characterized by high blood pressure and proteinuria and/or impaired liver and kidney function, pulmonary edema, hematological complications, seizures, or uteroplacental dysfunction (1,2). The prevalence of PE is 3-5% in the western world and it is a major contributor to maternal and fetal/neonatal morbidity (1). In places with limited access to healthcare, e.g. in large parts of Asia, Africa and South America, between one-tenth and one-quarter of maternal deaths are associated with hypertensive disorders of pregnancy (3). PE predisposes both mothers and offspring to a plethora of deleterious health outcomes. Mothers have increased risk of developing hypertension and other cardiovascular diseases (4), metabolic syndrome, and overweight and obesity years after pregnancy (5). Infants too are at risk of developing cardiovascular disease (6), metabolic syndrome and overweight and obesity later in life (6–8). The pathophysiological background of PE is elusive, but most likely heterogeneous, involving uterine-placental dysangiogenesis, endothelial dysfunction, immunological abnormalities, genetic factors, or metabolic disturbances (9).

Pregnancy requires major metabolic adaptations to ensure adequate nutrient supply for the fetus (10). An important aspect of this is changes in maternal insulin sensitivity (IS) and insulin resistance (IR) that is mediated by a range of maternal, placental, and fetal hormones (11). First trimester is characterized by a relative increase in insulin sensitivity, needed to ensure uptake of nutrients in the mother; this changes as insulin resistance develops during second-trimester (12,13).

Pre-pregnancy IR is associated with the development of gestational diabetes mellitus (GDM) (14), with ensuing risk of fetal over-nutrition and macrosomia (15–17) as well as PE (18). Early-onset PE is frequently associated with fetal growth restriction (FGR) (19), while late-onset PE is more often associated with macrosomia (2). Both fetal under- and over nutrition may lead to changes in body composition in the fetus that persist after birth (20,21). These, in turn, may lead to permanent metabolic alterations, epigenetic changes, and fetal programming, that ultimately result in the development of metabolic syndrome and its associated diseases later in life (6–8,22).

The risk of developing PE increases with increasing body mass index (BMI) (23). Furthermore, adiponectin and leptin play a major role in the metabolic adaptations to pregnancy. There is a small placental contribution to maternal leptin levels (24,25), but not to the extent that it obviates the relation between maternal BMI and leptin. A number of hormonal factors regulate the insulin signaling pathway and control the balance between IS and IR in order to accommodate pregnancy (11) as well as compensate for the metabolic consequences of overweight and obesity (26). Several hormones have been suggested to be involved in first-trimester fetal growth, and placental growth hormone appears to be of particular significance (27). In pregnant women with BMI values on the extreme ends of the scale, insufficient homeostatic compensatory mechanisms may result in presentation of disease symptoms. This process may be reflected in adipocytokine levels, already in first-trimester (26).

Adiponectin and leptin are involved in a wide range of physiological processes and seem perturbed in GDM and PE (14,28–35). Serum adiponectin-leptin ratio (A/L ratio) is a surrogate marker of IS that correlates well with signs of metabolic syndrome in childhood and adolescence (36,37). It has been shown to correlate with insulin resistance in pregnancy (38). However, it has not been associated with adverse pregnancy outcomes.

In this study, we use A/L ratio as a marker of IS and hypothesize that A/L ratio is perturbed in the first-trimester of pregnancies that later develop PE. Furthermore, we assess the significance of the A/L ratio for fetal growth, as maternal, placental, and fetal leptin have been reported to contribute to intrauterine growth (24). Accordingly, this study examines the relationship between first-trimester A/L ratio and PE as well as the relationship between adiponectin, leptin, and A/L ratio and birth weight.

## Materials and methods

### Study design and participants

This study is a case-control sub-study of the 1997 to 2001 Copenhagen First-Trimester Screening Study focusing on screening for chromosomal disorders (39). Several sub-studies on fetal growth (27), PE (25,40–42), and sources of Down syndrome serum marker variability (43,44) have been performed. The adipocytokine data used were collected from 2003 to 2010 and stored in an anonymized database. Only singleton pregnancies with first-trimester blood samples (gestational age (GA) 10^+3^-13^+6^) were included. GA was determined by crown rump length (CRL). Selection of cases and controls is described in Laigaard et al (40). Briefly, 126 pregnancies that developed PE were selected for the study and 297 controls were matched with respect to maternal age, parity, and GA at time of sampling (40). Of the 126 women with PE, 98 had mild PE, 21 had severe PE, and seven had HELLP syndrome. Ten pregnancies ended in delivery before GA 34^+0^. Regarding ethnicity, 93% of the women were of North European descent. Demographic and clinical parameters are given in table 1.

**Table 1.**
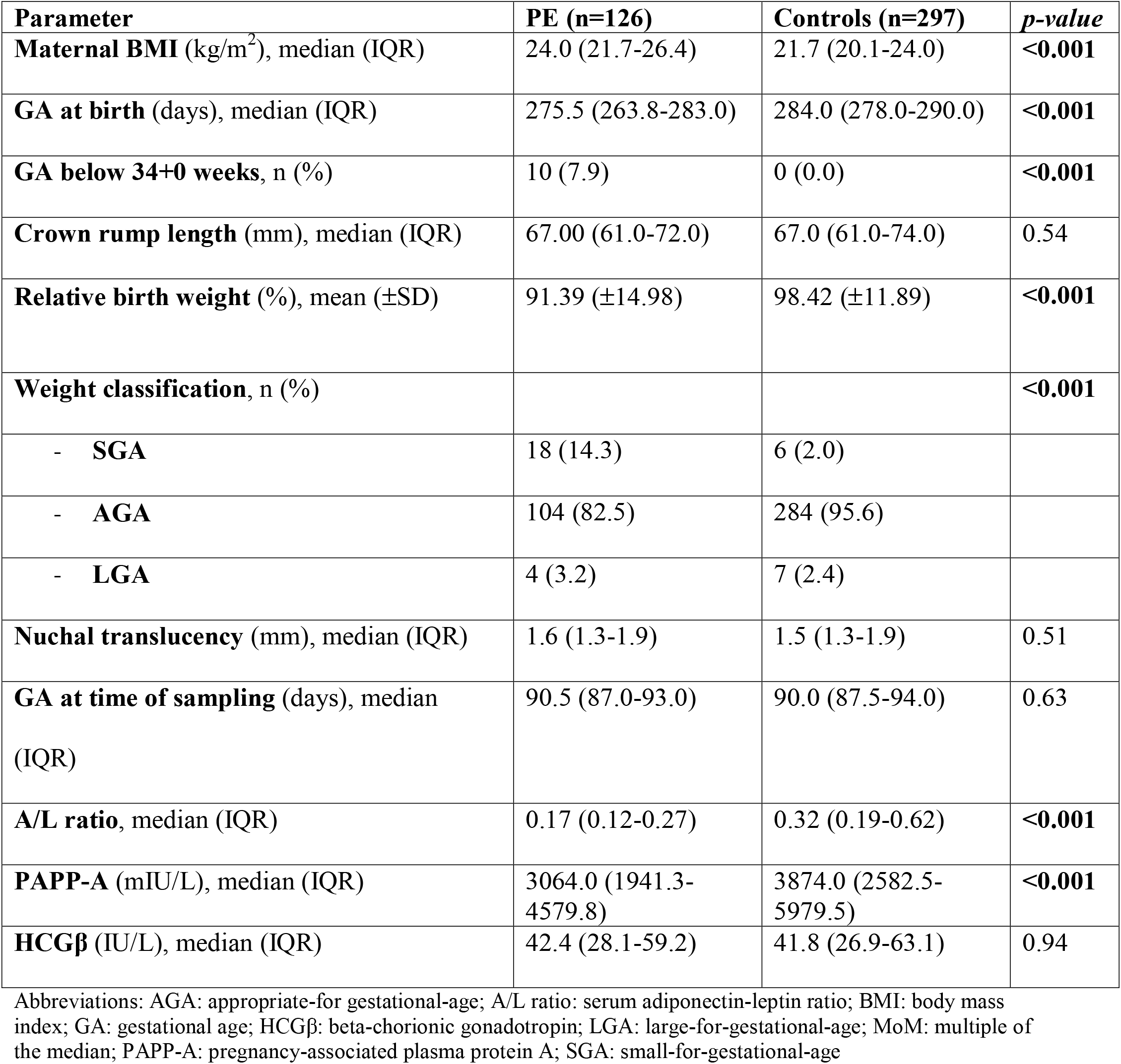
Demographic and clinical characteristics of PE and control pregnancies

### Data collection

Detailed clinical information, as well as pregnancy-associated plasma protein A (PAPP-A) and beta-chorionic gonadotropin (hCGβ) concentrations, were collected as part of the Copenhagen First-Trimester Screening Study, as previously described (39). PE pregnancies were categorized into one of three severity groups: mild PE, severe PE, and HELLP syndrome as per criteria from The International Society for the Study of Hypertension in Pregnancy (ISSHP) during the course of the First-Trimester Screening Study (45). Briefly, PE was defined as persistent hypertension, (either a systolic blood pressure ≥140 mm Hg or a diastolic blood pressure ≥90 mm Hg), occurring after 20 weeks of gestation in a previously normotensive woman, in combination with proteinuria, (≥0.3g in a 24-hour urine collection or dipstick urine analysis of ≥1+). Severe PE was defined by a diastolic blood pressure >110 mm Hg in combination with subjective symptoms and/or abnormal laboratory findings. HELLP syndrome was defined by the presence of hemolysis, elevated liver enzymes, and low platelets. Early PE was defined as a PE pregnancy that resulted in delivery prior to GA 34^+0^.

### Biochemical measurements

All blood samples were collected in dry containers and kept at 4° C for a maximum of 48 hours until storage at −20° C. Adiponectin and leptin concentrations were measured *in singlo* following appropriate sample dilution (25,26) using the Human Adiponectin Enzyme-Linked Immunosorbent Assay (ELISA) development kit, Duo Set (DY1065), and the Human Leptin ELISA development kit, Duo Set (DY398), R&D Systems, Minneapolis, USA. The functional detection limits of the assays were 62.5 pg/ml (for adiponectin) and 31.25 pg/ml (for leptin).

Evaluation of pre-analytic variables demonstrated that both adiponectin and leptin were stable for at least 48 hours at 23° C and 10 freeze-thaw cycles. The intra-assay and inter-assay coefficients of variation were <5% and <10%, respectively for adiponectin and leptin.

### Data analysis

Birth weights corrected for GA, relative birth weight, were calculated using intrauterine growth curves developed by Marsal et al. (46). Weight classifications small-for-gestational-age (SGA), appropriate-for-gestational-age (AGA), and large-for-gestational-age (LGA) were calculated using ± 24% as the upper and lower limits of relative birth weight (46). A/L ratio was calculated as ([adiponectin]/[leptin])/1000. Data were analyzed using generalized linear models following logarithmic transformation as appropriate. When necessary, non-parametric statistics were employed. A PAPP-A value corrected for GA at time of sampling was calculated using multiples of the median (MoM). MoM of logPAPP-A in both controls and PE pregnancies were calculated by performing a regression analysis in the control pregnancies of log_10_ PAPP-A on GA at time of sampling. All analyses were conducted using IBM SPSS Statistics, Version 28.0; Armonk, NY: IBM Corp.

### Ethics

The Copenhagen First Trimester Study was approved by the Scientific Ethics Committee for Copenhagen and Frederiksberg Counties (No. (KF) 01-288/97) and the Data Protection Agency approved the study protocol. All participants gave written informed consent.

## Results

### Comparison of PE and control pregnancies

A/L ratio was lower in PE pregnancies, 0.17 (IQR: 0.12 – 0.27) than in controls, 0.32 (IQR: 0.19 – 0.62) (p<0.001), (table 1). PAPP-A was significantly lower in PE pregnancies than controls, (p<0.001), (table 1). Furthermore, a significant association was found between A/L ratio and PAPP-A (log transformed and controlled for GA at time of testing) in PE (β = 1.432, 95% CI = 0.057 to 2.807). hCGβ concentrations were not significantly different between the two groups, (table 1). Offspring of women with PE had a lower GA at birth and lower relative birth weight compared to the control group, (p<0.001), (table 1). A larger proportion of PE pregnancies delivered SGA infants, 14.3% in PE vs. 2.0% in controls (p<0.001), (table 1). Compared to control pregnancies, women with PE had significantly higher BMI, (p<0.001), (table 1). CRL and nuchal translucency measurements did not differ significantly between PE pregnancies and controls, (table 1). 74 % of PE deliveries were induced compared to 16% in controls, (p<0.001).

A multiple logistic regression showed that PE was negatively associated with A/L ratio independent of maternal BMI (odds ratio = 0.082, 95% CI = 0.032 to 0.214). Adiponectin (area under the curve (AUC) = 0.632) and PAPP-A (AUC = 0.605) were negatively, and leptin (AUC = 0.712) was positively associated with PE. However, the A/L ratio was a better predictor of PE (AUC = 0.737), (figure 1).

**Figure 1.**
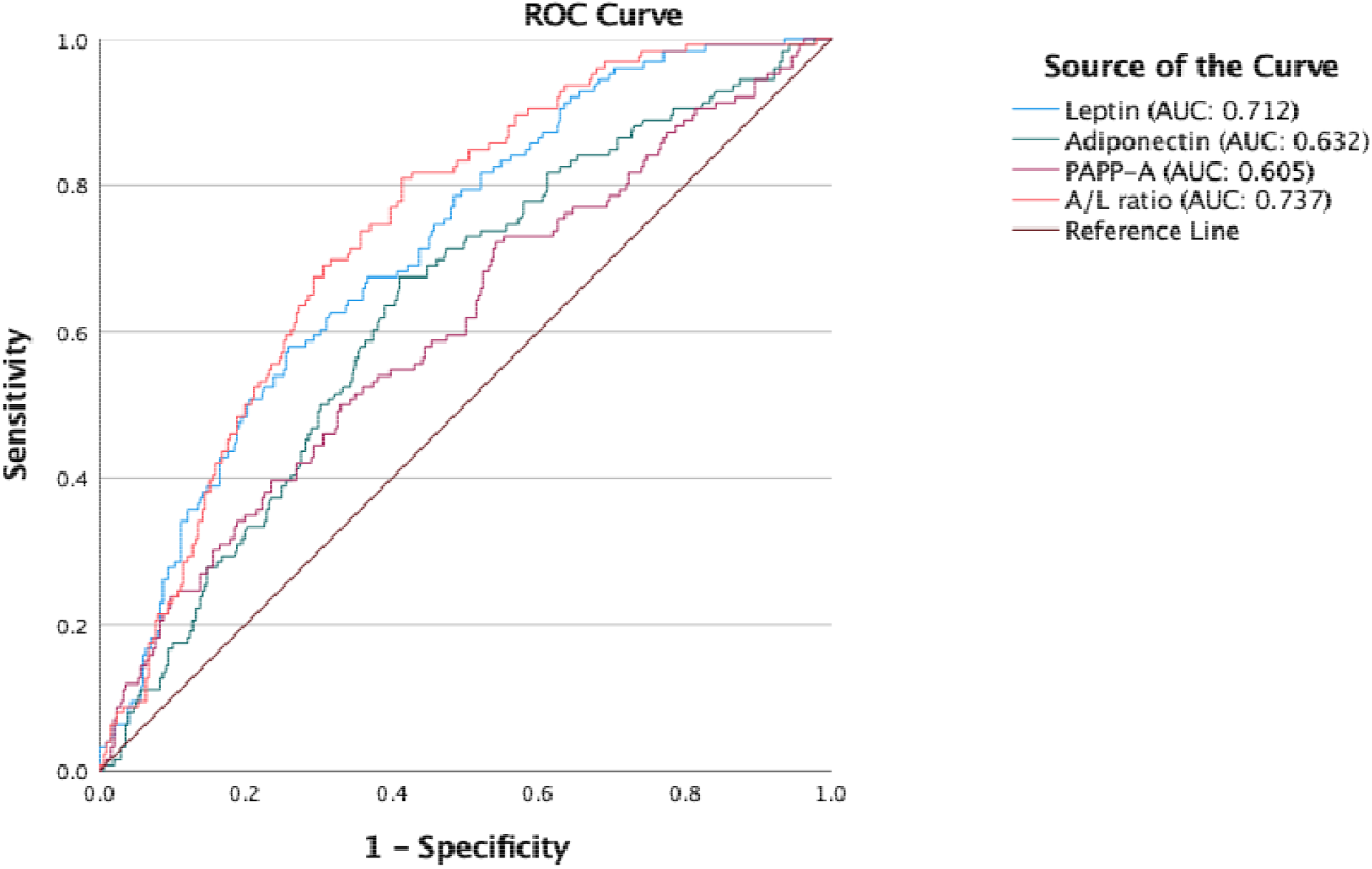
ROC curves of adiponectin, leptin, PAPP-A, and A/L ratio. A/L ratio was a better predictor of PE compared to adiponectin, leptin, and PAPP-A.

### Comparison of mild PE, severe PE, and HELLP syndrome

The demographic, clinical, and paraclinical characteristics of mild PE, severe PE, and HELLP pregnancies are presented in table 2. There was no discernable difference between A/L ratios in the different severity groups (figure 2). The proportion of early PE (<GA week 34) increased with increasing severity of PE, (p<0.001). There was no significant difference in relative birth weight between severity groups. However, the proportion of SGA infants was greater in severe PE and HELLP, (p = 0.006).

**Table 2.**
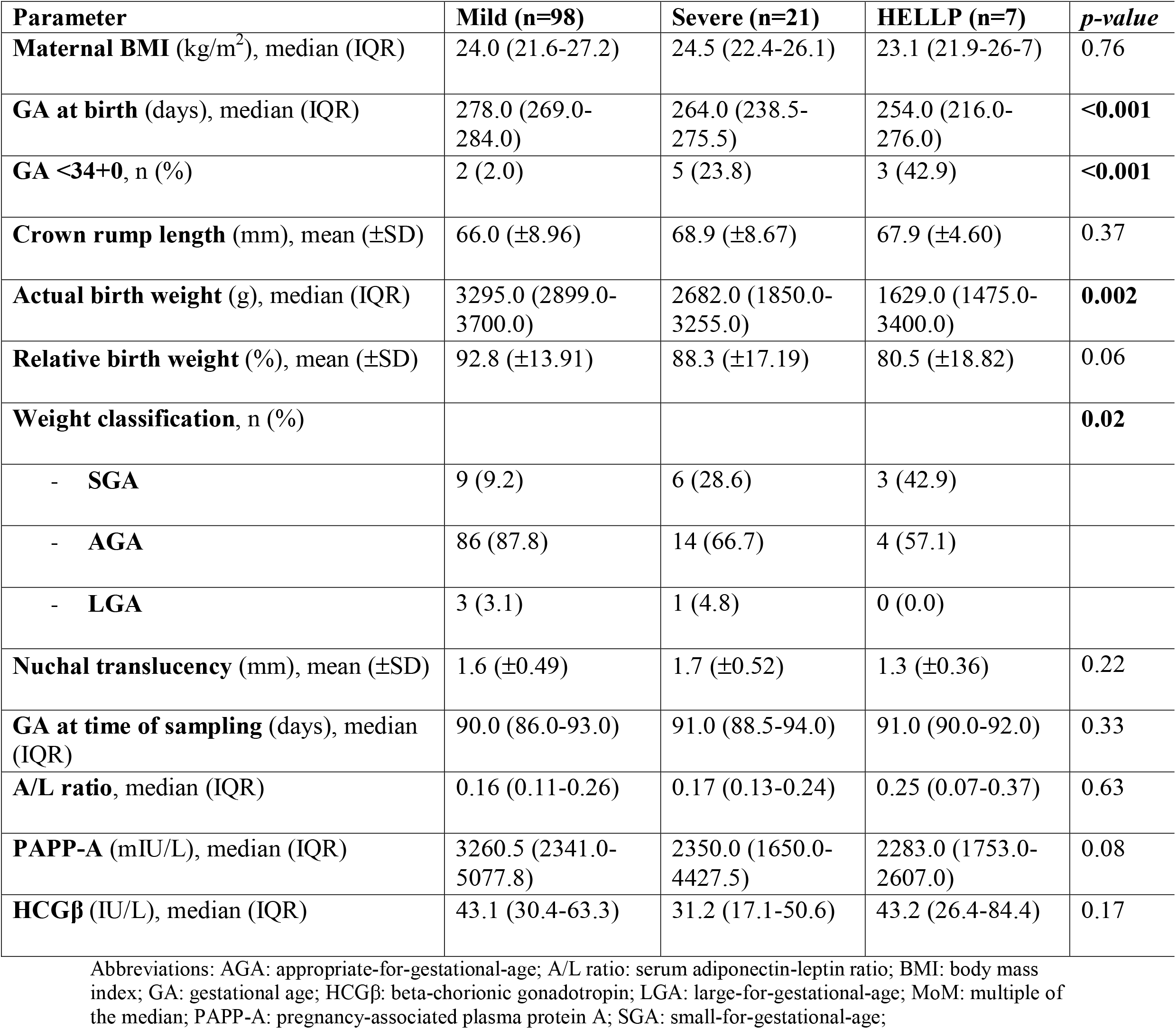
Demographic, clinical, and paraclinical characteristics of mild PE, severe PE and HELLP pregnancies

| Parameter | Mild (n=98) | Severe (n=21) | HELLP (n=7) | *p-value* |
| --- | --- | --- | --- | --- |
| **Maternal BMI** (kg/m<sup>2</sup>), median (IQR) | 24.0 (21.6-27.2) | 24.5 (22.4-26.1) | 23.1 (21.9-26-7) | 0.76 |
| **GA at birth** (days), median (IQR) | 278.0 (269.0-284.0) | 264.0 (238.5-275.5) | 254.0 (216.0-276.0) | **<0.001** |
| **GA <34+0**, n (%) | 2 (2.0) | 5 (23.8) | 3 (42.9) | **<0.001** |
| **Crown rump length** (mm), mean (±SD) | 66.0 (±8.96) | 68.9 (±8.67) | 67.9 (±4.60) | 0.37 |
| **Actual birth weight** (g), median (IQR) | 3295.0 (2899.0-3700.0) | 2682.0 (1850.0-3255.0) | 1629.0 (1475.0-3400.0) | **0.002** |
| **Relative birth weight** (%), mean (±SD) | 92.8 (±13.91) | 88.3 (±17.19) | 80.5 (±18.82) | 0.06 |
| **Weight classification**, n (%) |  |  |  | **0.02** |
| - **SGA** | 9 (9.2) | 6 (28.6) | 3 (42.9) |  |
| - **AGA** | 86 (87.8) | 14 (66.7) | 4 (57.1) |  |
| - **LGA** | 3 (3.1) | 1 (4.8) | 0 (0.0) |  |
| **Nuchal translucency** (mm), mean (±SD) | 1.6 (±0.49) | 1.7 (±0.52) | 1.3 (±0.36) | 0.22 |
| **GA at time of sampling** (days), median (IQR) | 90.0 (86.0-93.0) | 91.0 (88.5-94.0) | 91.0 (90.0-92.0) | 0.33 |
| **A/L ratio**, median (IQR) | 0.16 (0.11-0.26) | 0.17 (0.13-0.24) | 0.25 (0.07-0.37) | 0.63 |
| **PAPP-A** (mIU/L), median (IQR) | 3260.5 (2341.0-5077.8) | 2350.0 (1650.0-4427.5) | 2283.0 (1753.0-2607.0) | 0.08 |
| **HCGβ** (IU/L), median (IQR) | 43.1 (30.4-63.3) | 31.2 (17.1-50.6) | 43.2 (26.4-84.4) | 0.17 |

Abbreviations: AGA: appropriate-for-gestational-age; A/L ratio: serum adiponectin-leptin ratio; BMI: body mass index; GA: gestational age; HCGβ: beta-chorionic gonadotropin; LGA: large-for-gestational-age; MoM: multiple of the median; PAPP-A: pregnancy-associated plasma protein A; SGA: small-for-gestational-age;

**Figure 2.**
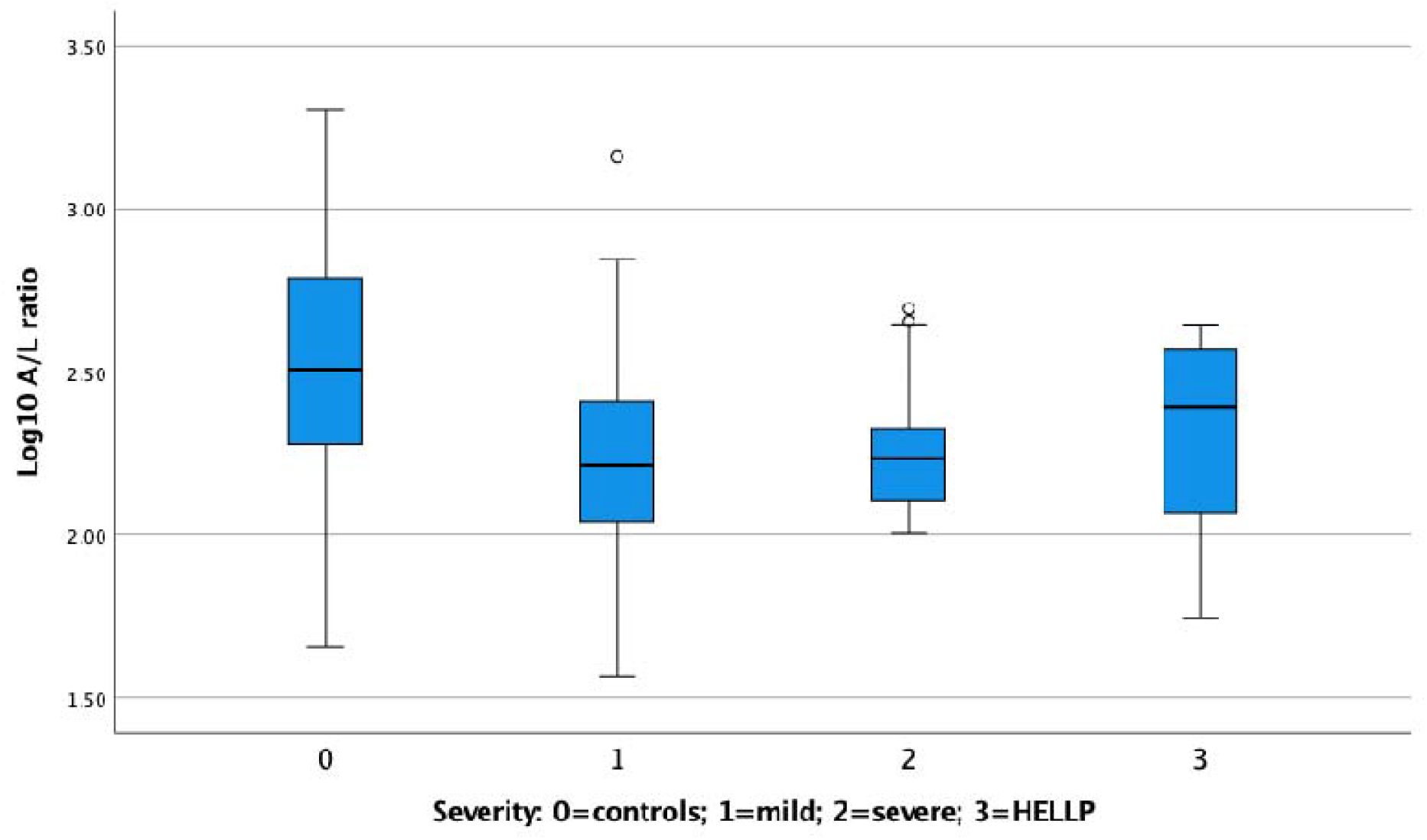
Box-plot of A/L ratio in controls, mild PE, severe PE, and HELLP. There was no discernable difference between A/L ratios in the different severity groups.

### Comparison of early and late PE

Demographic, clinical, and paraclinical characteristics of early and late PE are presented in table 3. No significant difference in A/L ratio between early and late PE was found. More frequently early PE pregnancies were classified as severe PE and HELLP syndrome compared with late PE pregnancies, 80.0% and 17.2%, respectively, (p<0.001). Early PE infants had a significantly lower relative birth weight than late PE infants, (p<0.001). There was a difference in size for GA between early and late PE, where 80.0% were SGA in early PE and 8.6% were SGA in late PE, (p<0.001).

**Table 3.** Demographic, clinical, and paraclinical characteristics of early and late PE

| **Parameter** | **Early PE (n=10)** | **Late PE (n=116)** | ***p*-value** |
| --- | --- | --- | --- |
| **Maternal BMI** (kg/m<sup>2</sup>), median (IQR) | 25.6 (23.0-28.8) | 24.0 (21.6-26.3) | 0.20 |
| **GA at birth** (days), median (IQR) | 210.5 (201.0-229.8) | 276.0 (268.0-283.8) | **<0.001** |
| **PE severity**, n (%) |  |  | **<0.001** |
| - **Mild** | 2 (20.0) | 96 (82.8) |  |
| - **Severe** | 5 (50.0) | 16 (13.8) |  |
| - **HELLP** | 3 (30.0) | 4 (3.4) |  |
| **Crown rump length** (mm), median (IQR) | 69.5 (62.3-72.3) | 67.0 (60.3-72.0) | 0.41 |
| **Relative birth weight** (%), mean (±SD) | 70.2 (±6.79) | 93.2 (±14.07) | **<0.001** |
| **Weight classification**, n (%) |  |  | **<0.001** |
| - **SGA** | 8 (80.0) | 10 (8.6) |  |
| - **AGA** | 2 (20.0) | 102 (87.9) |  |
| - **LGA** | 0 (0.0) | 4 (3.4) |  |
| **Nuchal translucency** (mm), median (IQR) | 1.7 (1.4-1.8) | 1.5 (1.3-1.9) | 0.98 |
| **GA at time of sampling** (days), median (IQR) | 90.5 (87.5-93.0) | 90.5 (87.0-93.0) | 0.85 |
| **A/L ratio**, median (IQR) | 0.13 (0.11-0.25) | 0.17 (0.12-0.27) | 0.45 |
| **PAPP-A** (mIU/L), median (IQR) | 1761.0 (1470.0-2194.3) | 3239.5 (2291.5-5019.8) | **0.003** |
| **HCGβ** (IU/L), median (IQR) | 42.4 (28.1-59.2) | 42.4 (29.6-61.9) | 0.21 |

Abbreviations: AGA: appropriate for gestational age; A/L ratio: serum adiponectin-leptin ratio; BMI: body mass index; GA: gestational age; HCGβ : beta-chorionic gonadotropin; LGA: large for gestational age; MoM: multiple of the median; PAPP-A: pregnancy-associated plasma protein A; PE: pre-eclampsia; SGA: small for gestational age

### Relative birth weight and A/L ratio

A significant negative correlation was found between relative birth weight and A/L ratio in controls but not in PE pregnancies, (*β* = −0.144, CI = −9.944 to −0.093). This was independent of maternal BMI. The relationship was valid also when only considering AGA controls (*β* = −0.189, 95% CI = −10.370 to −1.439), again independent of maternal BMI. After correction for maternal BMI, leptin was significantly associated with relative birth weight, (*β* = 0.197, 95% CI = 2.361 to 14.353), while adiponectin was not significantly associated. No association was found between A/L ratio and CRL in either PE or control pregnancies, independent of maternal BMI.

## Discussion

In this study we showed, for the first time, that A/L ratio in first-trimester was significantly lower in pregnancies that developed PE compared to controls. Previous studies have either not examined first-trimester pregnancies (38,47,48) or examined the effect of A/L ratio in first-trimester obese women who developed PE (26). In the latter study, a decrease in A/L ratio, albeit insignificant, was found (26). A/L ratio was, however, associated with development of GDM in the same cohort (49). This is compatible with the inverse correlation between A/L ratio and the homeostasis model assessment of IR in pregnancy (38). The A/L ratio decreased with maternal BMI, a known risk factor of PE (26), but the A/L ratio outperformed maternal BMI as a predictor of PE.

The leptin/adiponectin ratio (a marker of insulin resistance) is associated with cardiometabolic risk factors in children (36,37). Furthermore, in adults, a decreased A/L ratio has been shown to be associated with IR in various metabolic disorders, including diabetes mellitus (50,51), polycystic ovary syndrome (PCOS) (52–54), metabolic syndrome (55), atherosclerosis (56) and obesity (57). The reduction in A/L ratio, thus, likely reflects increasing IR. This could also explain the association between PE and PCOS (58).

In first-trimester PE pregnancies, the change in A/L ratio is driven by both a decrease in adiponectin and an increase in leptin. Adiponectin is a protective factor against cardiometabolic diseases (59), so the association between PE and later hypertension and diabetes type 2 could be explained by this relation. Adiponectin is not synthesized by the placenta, so the changes in adiponectin concentrations are the result of physiological changes in the mother. Leptin is partly synthesized by the placenta and largely by adipose tissue in the mother (33). However, the effect of leptin is modulated by the presence of soluble leptin receptor as well as variable expression of leptin receptors in different tissues (60). As pregnancy requires metabolic adaptations through gestation, mediated by several endocrine axes, the present study precludes a more precise analysis of the causation of the changes in adiponectin and leptin.

The receiver operating characteristic (ROC) curve analysis showed that A/L ratio was a better predictor of PE than leptin and adiponectin individually. However, the AUC of A/L ratio is so small that it precludes a use of A/L ratio as a first-trimester single marker of later PE. Likewise, PAPP-A is a marker for PE, but only used as a single marker in high risk pregnancies. However, it is possible that A/L ratio might contribute to improving the efficiency and clinical utility of either PE screening algorithms based on clinical information (61) or algorithms including ultrasound and biochemical markers (62). An advantage of a biochemical marker that may be measured on dried filter paper blood spots is that it may be used in populations with challenged access to prenatal care. This might be of particular importance in low- and middle-income countries.

There was no discernable difference between A/L ratios in the different severity groups of PE or between early and late PE. Thus, the A/L ratio in first trimester cannot be used to discern severity or early delivery in PE pregnancies. This suggests that IR predisposes to PE but other factors determine clinical presentation. A significantly larger cohort of PE cases will be needed to further investigate the performance of A/L ratio in early- and late-onset PE, severe PE and HELLP. Samples that were few in number in the present study.

A significant positive association between PAPP-A - a known marker for PE, placental dysfunction, and FGR - and A/L ratio was found in PE pregnancies. This is interesting as the growth hormone – insulin-like growth factor (IGF) axis, where PAPP-A is a placenta-derived insulin-like growth factor binding protein (IGFBP) −2, −4, and −5 protease (63), plays a major role for both fetal and placental growth (possibly due to its importance for bioavailability of IGF) (64). This specific finding needs to be confirmed through replication in other data sets.

A negative relationship between A/L ratio and relative birth weight was only found in controls, suggesting that the homeostatic mechanisms involving adiponectin and leptin are perturbed in PE. This finding was independent of maternal BMI. One explanation could, however, be that 74% of PE pregnancies were induced, in contrast to 16% of controls. Thus, the timing of parturition is for the majority of PE cases determined by the obstetrician, and not by endocrine effector systems. The relationship between A/L ratio and relative birth weight was also apparent when examining just AGA controls, indicating that it is not the extreme relative birth weights driving the overall association. Interestingly, the association between A/L ratio and relative birth weight in controls was driven by a positive association between leptin and relative birth weight. Adiponectin did not contribute to this association. Thus, relative birth weight does not seem to be dependent on IS but rather on leptin associated pathways (24).

Leptin contributed independently to the negative association between A/L ratio and relative birth weight in controls. Leptin has a permissive role in several reproductive functions (65), and in the current context, one might hypothesize that leptin reflects whether maternal energy stores are sufficient sustain normal fetal growth. We know that the relationship between leptin and maternal BMI is perturbed at high BMI levels (26), and increased IR originating from adipose tissue can be a contributing factor (66).

PE is – in some cases - associated with reduced fetal size and as expected, we found that women with PE had infants with significantly lower relative birth weight compared with controls. Yet, interestingly, there was a lack of association between birth weight and A/L ratio in PE pregnancies. These findings suggest that disturbances in metabolic homeostasis are already apparent long before the development of PE symptoms, by definition in week 20, and have negative consequences for fetal growth. Alternatively, the above mentioned large difference in induction rates between PE pregnancies and controls may override the normal homeostatic mechanisms controlling time of parturition. However, the period of relative IS and optimal nutrient access in first-trimester may be reduced in women who develop PE as they instead risk experiencing premature and/or increased IR. This possible shift between IS and IR already in first-trimester may herald the PE associated impaired fetal growth.

Even though the study included a broad range of clinical parameters, some important variables, such as gender of offspring and gestational weight gain during pregnancy were not included. Another limitation of this study was that only total adiponectin levels were measured. Three forms of adiponectin, low-, medium-, and high-molecular-weight (HMW) adiponectin have been identified, where only HMW adiponectin has been shown to be associated to PE (67).

## Conclusions

A reduced A/L ratio is a first-trimester characteristic of pregnancies that later develop PE. However, neither severity nor time of onset was associated with A/L ratio. The A/L ratio, reflecting IS, is a better discriminator between PE and controls than either leptin or adiponectin. Relative birth weight was associated with A/L ratio, however, this was driven by leptin alone. Changes in birth weight do not seem to be dependent on insulin sensitivity, as is the case for PE, but rather on leptin associated pathways.

## Supporting information

STROBE Checklist

## Data Availability

All data produced in the present work are contained in the manuscript.

## List of abbreviations

AGA: appropriate-for-gestational-age
A/L ratio: serum adiponectin-leptin ratio
AUC: area under the curve
BMI: body mass index
CRL: crown rump length
ELISA: enzyme-linked immunosorbent assay
FGR: fetal growth restriction
GA: gestational age
GDM: gestational diabetes mellitus
HCGβ: beta-chorionic gonadotropin
HMW: high molecular weight
IGF: insulin-like growth factor
IGFBP: insulin-like growth factor binding protein
IR: insulin resistance
IS: insulin sensitivity
ISSHP: The International Society for the Study of Hypertension
LGA: large-for-gestational-age
MoM: multiple of the median
PAPP-A: pregnancy-associated plasma protein A
PCOS: polycystic ovary syndrome
PE: pre-eclampsia
ROC: receiver operating characteristic
SGA: small-for-gestational-age

## Declarations

### Ethics approval and consent to participate

The study was a sub-study of the Copenhagen First-Trimester Screening Study, which was approved by the Scientific Ethics Committee for Copenhagen and Frederiksberg Counties (No. (KF) 01-288/97). All participants gave written informed consent.

### Consent for publication

Not applicable.

### Availability of data and materials

The datasets generated and/or analyzed during the current study are not publicly available due to restrictions in handling personal data, and particularly because participants were recruited at a time when it was not standard practice to make samples/data publicly available but are available from the corresponding author on reasonable request.

### Authors’ contributions

VK, MC, PH, and UL conceived and designed the sub-study; SP, KW, ACS, LR, KS, and AT collected the data and performed clinical assessments; VK, MC, and PH did the data analysis; VK, MC, PH, UL, AKE, LR, AT, and JK interpreted the results of the data; VK, MC, PH, and UL drafted the work; all authors revised the work critically for intellectual content; all authors approved the final version of the work to be published; all authors are accountable for all aspects of the work in ensuring that questions related to the accuracy or integrity of any part of the work are appropriately investigated and resolved.

## Acknowledgements

We gratefully acknowledge the expert technical assistance of Pia Lind and Pernilla Rasmussen. We also gratefully acknowledge the financial support to the Copenhagen First Trimester Study from the Danish Medical Research Council, Copenhagen University, The John and Birthe Meyer Foundation, The Ivan Nielsen Foundation, The Else and Mogens Wedell-Wedellsborg Foundation, The Dagmar Marshall Foundation, The Egmont Foundation, The Fetal Medicine Foundation, The Augustinus Foundation, The Gangsted Foundation, The A.P. Møller Foundation, The Mads Clausens Foundation, The Copenhagen Hospital Corporation, and SAFE Network of Excellence. This research has been conducted using the Danish National Biobank resource, funded by the Novo Nordisk Foundation.

## References

1. Mol BWJ, Roberts CT, Thangaratinam S, Magee LA, de Groot CJM, Hofmeyr GJ. Pre-eclampsia. Lancet. 2016 Mar 5;387(10022):999–1011.

2. Magee LA, Nicolaides KH, von Dadelszen P. Preeclampsia. N Engl J Med. 2022 May 12;386(19):1817–32.

3. WHO Recommendations for Prevention and Treatment of Pre-Eclampsia and Eclampsia [Internet]. Geneva: World Health Organization; 2011 [cited 2020 Jun 3]. (WHO Guidelines Approved by the Guidelines Review Committee). Available from: http://www.ncbi.nlm.nih.gov/books/NBK140561/

4. Hauspurg A, Countouris ME, Catov JM. Hypertensive Disorders of Pregnancy and Future Maternal Health: How Can the Evidence Guide Postpartum Management? Curr Hypertens Rep. 2019 Nov 27;21(12):96.

5. Alonso-Ventura V, Li Y, Pasupuleti V, Roman YM, Hernandez AV, Pérez-López FR. Effects of preeclampsia and eclampsia on maternal metabolic and biochemical outcomes in later life: a systematic review and meta-analysis. Metab Clin Exp. 2020 Jan;102:154012.

6. Karatza AA, Dimitriou G. Preeclampsia emerging as a novel risk factor for cardiovascular disease in the offspring. Curr Pediatr Rev. 2019 Dec 23;

7. Goffin SM, Derraik JGB, Groom KM, Cutfield WS. Maternal pre-eclampsia and long-term offspring health: Is there a shadow cast? Pregnancy Hypertens. 2018 Apr;12:11–5.

8. Pinheiro TV, Brunetto S, Ramos JGL, Bernardi JR, Goldani MZ. Hypertensive disorders during pregnancy and health outcomes in the offspring: a systematic review. J Dev Orig Health Dis. 2016;7(4):391–407.

9. Burton GJ, Redman CW, Roberts JM, Moffett A. Pre-eclampsia: pathophysiology and clinical implications. BMJ. 2019 15;366:2381.

10. Mouzon SH de Lassance L. Endocrine and metabolic adaptations to pregnancy; impact of obesity. Horm Mol Biol Clin Investig. 2015 Oct;24(1):65–72.

11. Sonagra AD, Biradar SM, k D, Murthy D S J. Normal pregnancy-a state of insulin resistance. J Clin Diagn Res. 2014 Nov;8(11):CC01–03.

12. Catalano PM, Roman-Drago NM, Amini SB, Sims EA. Longitudinal changes in body composition and energy balance in lean women with normal and abnormal glucose tolerance during pregnancy. Am J Obstet Gynecol. 1998 Jul;179(1):156–65.

13. García-Patterson A, Gich I, Amini SB, Catalano PM, de Leiva A, Corcoy R. Insulin requirements throughout pregnancy in women with type 1 diabetes mellitus: three changes of direction. Diabetologia. 2010 Mar;53(3):446–51.

14. Plows JF, Stanley JL, Baker PN, Reynolds CM, Vickers MH. The Pathophysiology of Gestational Diabetes Mellitus. Int J Mol Sci. 2018 Oct 26;19(11).

15. Brunner S, Schmid D, Hüttinger K, Much D, Heimberg E, Sedlmeier EM, et al. Maternal insulin resistance, triglycerides and cord blood insulin in relation to post-natal weight trajectories and body composition in the offspring up to 2 years. Diabet Med. 2013 Dec;30(12):1500–7.

16. Crume TL, Shapiro AL, Brinton JT, Glueck DH, Martinez M, Kohn M, et al. Maternal fuels and metabolic measures during pregnancy and neonatal body composition: the healthy start study. J Clin Endocrinol Metab. 2015 Apr;100(4):1672–80.

17. Shapiro ALB, Schmiege SJ, Brinton JT, Glueck D, Crume TL, Friedman JE, et al. Testing the fuel-mediated hypothesis: maternal insulin resistance and glucose mediate the association between maternal and neonatal adiposity, the Healthy Start study. Diabetologia. 2015 May;58(5):937–41.

18. Cho GJ, Park JH, Shin SA, Oh MJ, Seo HS. Metabolic syndrome in the non-pregnant state is associated with the development of preeclampsia. Int J Cardiol. 2016 Jan 15;203:982–6.

19. Audette MC, Kingdom JC. Screening for fetal growth restriction and placental insufficiency. Semin Fetal Neonatal Med. 2018;23(2):119–25.

20. Dessì A, Puddu M, Ottonello G, Fanos V. Metabolomics and fetal-neonatal nutrition: between “not enough” and “too much.” Molecules. 2013 Sep 25;18(10):11724–32.

21. Hales CN, Barker DJ. The thrifty phenotype hypothesis. Br Med Bull. 2001;60:5–20.

22. Maiorana A, Del Bianco C, Cianfarani S. Adipose Tissue: A Metabolic Regulator. Potential Implications for the Metabolic Outcome of Subjects Born Small for Gestational Age (SGA). Rev Diabet Stud. 2007;4(3):134–46.

23. Motedayen M, Rafiei M, Rezaei Tavirani M, Sayehmiri K, Dousti M. The relationship between body mass index and preeclampsia: A systematic review and meta-analysis. Int J Reprod Biomed (Yazd). 2019 Jul;17(7):463–72.

24. de Knegt VE, Hedley PL, Kanters JK, Thagaard IN, Krebs L, Christiansen M, et al. The Role of Leptin in Fetal Growth during Pre-Eclampsia. Int J Mol Sci. 2021 Apr 27;22(9).

25. Hedley PL, Placing S, Wøjdemann K, Carlsen AL, Shalmi AC, Sundberg K, et al. Free leptin index and PAPP-A: a first trimester maternal serum screening test for pre-eclampsia. Prenat Diagn. 2010 Feb;30(2):103–9.

26. Thagaard IN, Hedley PL, Holm JC, Lange T, Larsen T, Krebs L, et al. Leptin and Adiponectin as markers for preeclampsia in obese pregnant women, a cohort study. Pregnancy Hypertens. 2019 Jan;15:78–83.

27. Pedersen NG, Juul A, Christiansen M, Wøjdemann KR, Tabor A. Maternal serum placental growth hormone, but not human placental lactogen or insulin growth factor-1, is positively associated with fetal growth in the first half of pregnancy. Ultrasound Obstet Gynecol. 2010 Nov;36(5):534–41.

28. Retnakaran A, Retnakaran R. Adiponectin in pregnancy: implications for health and disease. Curr Med Chem 2012;19(32):5444–50.

29. Herrid M, Palanisamy SKA, Ciller UA, Fan R, Moens P, Smart NA, et al. An updated view of leptin on implantation and pregnancy: a review. Physiol Res. 2014;63(5):543–57.

30. Miehle K, Stepan H, Fasshauer M. Leptin, adiponectin and other adipokines in gestational diabetes mellitus and pre-eclampsia. Clin Endocrinol (Oxf). 2012 Jan;76(1):2–11.

31. Dos Santos E, Duval F, Vialard F, Dieudonné MN. The roles of leptin and adiponectin at the fetal-maternal interface in humans. Horm Mol Biol Clin Investig. 2015 Oct;24(1):47–63.

32. Hauguel-de Mouzon S, Lepercq J, Catalano P. The known and unknown of leptin in pregnancy. Am J Obstet Gynecol. 2006 Jun;194(6):1537–45.

33. D’Ippolito S, Tersigni C, Scambia G, Di Simone N. Adipokines, an adipose tissue and placental product with biological functions during pregnancy. Biofactors. 2012 Feb;38(1):14–23.

34. Pérez-Pérez A, Toro A, Vilariño-García T, Maymó J, Guadix P, Dueñas JL, et al. Leptin action in normal and pathological pregnancies. J Cell Mol Med. 2018;22(2):716–27.

35. Dalamaga M, Srinivas SK, Elovitz MA, Chamberland J, Mantzoros CS. Serum adiponectin and leptin in relation to risk for preeclampsia: results from a large case-control study. Metab Clin Exp. 2011 Nov;60(11):1539–44.

36. Frithioff-Bøjsøe C, Lund MAV, Lausten-Thomsen U, Hedley PL, Pedersen O, Christiansen M, et al. Leptin, adiponectin, and their ratio as markers of insulin resistance and cardiometabolic risk in childhood obesity. Pediatr Diabetes. 2020 Mar;21(2):194–202.

37. Lausten-Thomsen U, Lund MAV, Frithioff-Bøjsøe C, Hedley PL, Pedersen O, Hansen T, et al. Reference values for leptin/adiponectin ratio in healthy children and adolescents. Clin Chim Acta. 2019 Jun;493:123–8.

38. Skvarca A, Tomazic M, Blagus R, Krhin B, Janez A. Adiponectin/leptin ratio and insulin resistance in pregnancy. J Int Med Res. 2013 Feb;41(1):123–8.

39. Wøjdemann KR, Shalmi AC, Christiansen M, Larsen SO, Sundberg K, Brocks V, et al. Improved first-trimester Down syndrome screening performance by lowering the false-positive rate: a prospective study of 9941 low-risk women. Ultrasound Obstet Gynecol. 2005 Mar;25(3):227–33.

40. Laigaard J, Sørensen T, Placing S, Holck P, Fröhlich C, Wøjdemann KR, et al. Reduction of the disintegrin and metalloprotease ADAM12 in preeclampsia. Obstet Gynecol. 2005 Jul;106(1):144–9.

41. De Villiers CP, Hedley PL, Placing S, Wøjdemann KR, Shalmi AC, Carlsen AL, et al. Placental protein-13 (PP13) in combination with PAPP-A and free leptin index (fLI) in first trimester maternal serum screening for severe and early preeclampsia. Clin Chem Lab Med. 2017 Nov 27;56(1):65–74.

42. Christiansen M, Hedley PL, Placing S, Wøjdemann KR, Carlsen AL, Jørgensen JM, et al. Maternal Serum Resistin Is Reduced in First Trimester Preeclampsia Pregnancies and Is a Marker of Clinical Severity. Hypertens Pregnancy. 2015 Nov;34(4):422–33.

43. Laigaard J, Larsen SO, Pedersen NG, Hedley PL, Gjerris AC, Wøjdemann KR, et al. ADAM 12-S in first trimester: fetal gender, smoking and maternal age influence the maternal serum concentration. Prenat Diagn. 2009 May;29(5):525–7.

44. Laigaard J, Pedersen NG, Larsen SO, Hedley PL, Wøjdemann K, Gjerris AC, et al. ADAM12 in first trimester maternal serum from pregnancies conceived by assisted reproduction techniques (ART). Prenat Diagn. 2009 Jun;29(6):628–9.

45. Brown MA, Lindheimer MD, de Swiet M, Van Assche A, Moutquin JM. The classification and diagnosis of the hypertensive disorders of pregnancy: statement from the International Society for the Study of Hypertension in Pregnancy (ISSHP). Hypertens Pregnancy. 2001;20(1):IX–XIV.

46. Marsál K, Persson PH, Larsen T, Lilja H, Selbing A, Sultan B. Intrauterine growth curves based on ultrasonically estimated foetal weights. Acta Paediatr. 1996 Jul;85(7):843–8.

47. Nakatsukasa H, Masuyama H, Takamoto N, Hiramatsu Y. Circulating leptin and angiogenic factors in preeclampsia patients. Endocr J. 2008 Jul;55(3):565–73.

48. Ouyang Y, Chen H, Chen H. Reduced plasma adiponectin and elevated leptin in pre-eclampsia. xInt J Gynaecol Obstet. 2007 Aug;98(2):110–4.

49. Thagaard IN, Krebs L, Holm JC, Lange T, Larsen T, Christiansen M. Adiponectin and leptin as first trimester markers for gestational diabetes mellitus: a cohort study. Clin Chem Lab Med. 2017 Oct 26;55(11):1805–12.

50. Zaletel J, Barlovic DP, Prezelj J. Adiponectin-leptin ratio: a useful estimate of insulin resistance in patients with Type 2 diabetes. J Endocrinol Invest. 2010 Sep;33(8):514–8.

51. Inoue M, Maehata E, Yano M, Taniyama M, Suzuki S. Correlation between the adiponectin-leptin ratio and parameters of insulin resistance in patients with type 2 diabetes. Metab Clin Exp. 2005 Mar;54(3):281–6.

52. Obirikorang C, Owiredu WKBA, Adu-Afram S, Acheampong E, Asamoah EA, Antwi-Boasiakoh EK, et al. Assessing the variability and predictability of adipokines (adiponectin, leptin, resistin and their ratios) in non-obese and obese women with anovulatory polycystic ovary syndrome. BMC Res Notes. 2019 Aug 15;12(1):513.

53. Sarray S, Madan S, Saleh LR, Mahmoud N, Almawi WY. Validity of adiponectin-to-leptin and adiponectin-to-resistin ratios as predictors of polycystic ovary syndrome. Fertil Steril. 2015 Aug;104(2):460–6.

54. Svendsen PF, Christiansen M, Hedley PL, Nilas L, Pedersen SB, Madsbad S. Adipose expression of adipocytokines in women with polycystic ovary syndrome. Fertil Steril. 2012 Jul;98(1):235–41.

55. Mirza S, Qu HQ, Li Q, Martinez PJ, Rentfro AR, McCormick JB, et al. Adiponectin/leptin ratio and metabolic syndrome in a Mexican American population. Clin Invest Med. 2011 Oct 1;34(5):E290.

56. Frühbeck G, Catalán V, Rodríguez A, Gómez-Ambrosi J. Adiponectin-leptin ratio: A promising index to estimate adipose tissue dysfunction. Relation with obesity-associated cardiometabolic risk. Adipocyte. 2018 02;7(1):57–62.

57. Frühbeck G, Catalán V, Rodríguez A, Ramírez B, Becerril S, Salvador J, et al. Adiponectin-leptin Ratio is a Functional Biomarker of Adipose Tissue Inflammation. Nutrients. 2019 Feb 22;11(2).

58. Naver KV, Grinsted J, Larsen SO, Hedley PL, Jørgensen FS, Christiansen M, et al. Increased risk of preterm delivery and pre-eclampsia in women with polycystic ovary syndrome and hyperandrogenaemia. BJOG. 2014 Apr;121(5):575–81.

59. Younus S, Rodgers G. Biomarkers associated with cardiometabolic risk in obesity. Am Heart Hosp J. 2011;9(1):E28-32.

60. Fasshauer M, Blüher M. Adipokines in health and disease. Trends Pharmacol Sci. 2015 Jul;36(7):461–70.

61. Gestational Hypertension and Preeclampsia: xACOG Practice Bulletin, Number 222. Obstet Gynecol. 2020 Jun;135(6):e237–60.

62. Chaemsaithong P, Sahota DS, Poon LC. First trimester preeclampsia screening and prediction. Am J Obstet Gynecol. 2022 Feb;226(2S):S1071-S1097.e2.

63. Steffensen LB, Conover CA, Oxvig C. PAPP-A and the IGF system in atherosclerosis: what’s up, what’s down? Am J Physiol Heart Circ Physiol. 2019 Nov 1;317(5):H1039–49.

64. Kaur H, Muhlhausler BS, Roberts CT, Gatford KL. The growth hormone-insulin like growth factor axis in pregnancy. J Endocrinol. 2021 Sep 1;JOE-21-0087.R1.

65. Childs GV, Odle AK, MacNicol MC, MacNicol AM. The Importance of Leptin to Reproduction. Endocrinology. 2021 Feb 1;162(2).

66. Spradley FT. Metabolic abnormalities and obesity’s impact on the risk for developing preeclampsia. Am J Physiol Regul Integr Comp Physiol. 2017 Jan 1;312(1):R5–12.

67. Takemura Y, Osuga Y, Koga K, Tajima T, Hirota Y, Hirata T, et al. Selective increase in high molecular weight adiponectin concentration in serum of women with preeclampsia. J Reprod Immunol. 2007 Feb;73(1):60–5.

